# Nutritional screening in mental health and learning disability inpatient services: Dietitians’ perspectives on practices, barriers and tool suitability

**DOI:** 10.64898/2026.08.25.26361293

**Authors:** Steph Smith, April Leong, Garreth Burke, Rebecca Guerin

## Abstract

**Introduction:** People with severe mental illness (SMI) and learning disabilities (LD) experience significant health inequalities, with diet-related conditions contributing substantially to early and preventable death. Despite high levels of nutritional risk, the presence and effectiveness of nutritional screening in mental health (MH) and LD settings remains under-researched. This study aimed to investigate nutritional screening practices in UK inpatient MH and LD services from the perspectives of dietitians.

**Methods:** A cross-sectional mixed-methods study was conducted using a novel 22-question online survey. Data was collected via the British Dietetic Association Mental Health Specialist Group (April-June 2025). Quantitative data was analysed descriptively and qualitative data by reflexive thematic analysis. Findings were integrated and presented thematically. Ethical approval was granted by Teesside University (2025Mar26544).

**Results:** Forty-seven dietitians participated, most with substantial dietetic experience, from a range of MH settings. Screening practices were widely established and supported by policy and audit. However, participants reported low confidence in screening translating into meaningful patient care. Barriers to screening included appropriateness of available tools, time constraints, difficulty engaging distressed patients and poor prioritisation of physical health. Digital integration and wider infrastructure were also important. Dietitians rarely undertook screening directly, instead holding secondary or leadership roles, while screening was most often completed by nursing staff who were often perceived to place limited importance on the process. Existing tools, particularly the Malnutrition Universal Screening Tool (MUST), were viewed as insufficiently capturing the broader nutritional risks relevant to MH/LD populations, leading some services to adopt bespoke, unvalidated tools.

**Conclusion:** Concerns regarding the suitability of existing nutritional screening tools in MH/LD settings are consistent with previous literature. However, we suggest cautious use of unvalidated bespoke tools. Whilst there was no clear front runner, MH specific tools such as the St Andrew’s Nutrition Screening Instrument (SANSI) and the NutriMental Screener warrant further evaluation. Importantly, findings indicate that optimising tool choice alone is unlikely to improve screening effectiveness. Nutritional screening must be embedded within clear care pathways, supported by organisational leadership, digital infrastructure, and multiprofessional engagement to move beyond procedural completion and support meaningful clinical action to improve patient care.

**Practitioner Points:**

- Nutritional screening in mental health and learning disability inpatient services is often experienced as procedural with limited patient impact unless clearly linked to care pathways and follow-up action.
- Screening tool choice alone is unlikely to improve practice. Organisational leadership, adequate time, digital integration, and multiprofessional engagement are critical, alongside the use of reasonable adjustments and pragmatic approaches where mental state or communication differences impact screening.
- Dietitians typically hold advisory or leadership roles rather than completing screening directly, highlighting the importance of engaging nursing staff and supporting multiprofessional ownership of nutritional screening processes.

## Background

### Nutritional Risk in Mental Health and Learning Disabilities

People with severe mental illness (SMI) and learning disabilities (LD; the term commonly used in the UK for intellectual disability) experience marked health inequalities shaped by multimorbidity (1), diagnostic overshadowing (2), communication barriers (3), fragmented care (4), and socioeconomic exclusion (5), dying on average 15–20 years earlier than the general population (6,7). A substantial proportion of these deaths are avoidable: 40.2% among people with LD and 60% among people with SMI (8,9), with diet-related conditions, including cardiovascular disease, contributing substantially to excess mortality (7,9). Obesity is common among people with SMI and reflects a complex interplay of biopsychosocial factors. These include medication-related changes in appetite, cravings, dietary disinhibition, emotional eating, and dietary intake and composition, although findings are inconsistent despite these experiences being described prominently by people with lived experience (10). Poor diet quality (11), low physical activity (12) and socioeconomic disadvantage (13) may further contribute to obesity risk. Prevalence is higher in inpatient settings, which are frequently described as obesogenic (14).

Undernutrition is also a concern in mental health (MH) settings, where prevalence exceeds that of general hospitals, community, care home and rehabilitation settings (15), and is associated with greater symptom severity and poorer functioning (16). In LD populations, undernutrition rates are also higher than in the general population (17).

People with LD are disproportionately affected by nutrition-related or nutrition-impacting conditions, including Type 1 diabetes (18), thyroid disorders (19), osteoporosis (20), constipation (21) and dysphagia (22) while conditions such as epilepsy can further impact nutritional management (23). In MH populations, gastrointestinal issues including constipation (24) are frequently reported. Disordered eating (25) and psychotic or obsessive-compulsive symptoms leading to acute food refusal or atypical eating patterns (26) are also common.

### Nutritional Screening

Nutritional screening enables rapid identification of risk and early intervention. In MH settings, where malnutrition is often overlooked (27) and access to dietetic care is limited (28), effective screening is essential for targeting scarce dietetic resource. However, few tools have been validated specifically for MH/LD populations (29). The Malnutrition Universal Screening Tool (MUST) (30) is widely used across health and care settings (31–33) and recommended in national clinical guidelines (34), but its ability to identify the breadth of nutritional risk n MH/LD populations has been questioned (35). The breadth of nutritional risk in MH/LD populations is reflected in a 2021 scoping review of nutritional screening, which encompassed under- and overnutrition alongside risks including constipation, dysphagia and disordered eating (29).

The St Andrew’s Nutrition Screening Instrument (SANSI) (35) was developed and initially validated in secure psychiatric settings to capture this broader risk profile, incorporating dysphagia, food and fluid refusal, and disordered eating alongside anthropometrics. Its use for initial risk stratification within nutritional care pathways has also been demonstrated (36).

The NutriMental Screener (37) and Approaches to Schizophrenia Communication Self Report (ASC-SR) checklist (38) have also been developed for MH settings. The NutriMental Screener has shown feasibility, and acceptability (39) though validation in UK MH/LD populations is lacking. A pilot study suggests the ASC-SR may support identification of nutrition-related medication side effects in United States SMI populations but further tool validation is needed (38). Existing tools, such as the Nutrition Risk Score, have also been adapted for MH populations (27), though validation of such modifications is limited.

**Table 1:** Comparison of key nutrition screening tools.

| **<u>Screening tool</u>** | **<u>Primary purpose</u>** | **<u>Items and structure</u>** | **<u>Key domains assessed</u>** | **<u>Key limitations and considerations</u>** |
| --- | --- | --- | --- | --- |
| **Malnutrition Universal Screening Tool (MUST) (30)** | Identifies adults with or at risk of malnutrition (undernutrition); also identifies obesity through BMI classification. | 3 items scored within a five-step screening and management process. | Body Mass Index (BMI), unintentional weight loss, acute disease effect (little or no nutritional intake secondary to acute illness). | Widely established across general adult healthcare settings but not developed specifically for mental health (MH) or learning disability (LD) populations. Focuses primarily on undernutrition and may not capture broader MH-related nutritional risks such as dysphagia, disordered eating or food refusal, unless accompanied by low weight and/or significant weight loss. |
| **St Andrew's Nutrition Screening Instrument (SANSI) (35)** | Developed to identify nutritional risk in adolescent and adult secure psychiatric inpatient settings. Identifies a range | 12 risk items within a four-step instrument: BMI, weight change and 10 yes/no questions | BMI, weight change, dysphagia, food refusal, fluid refusal, disordered eating and other | Designed specifically for MH/LD populations, although evidence base is limited compared to more established |

| <u>Screening tool</u> | <u>Primary purpose</u> | <u>Items and structure</u> | <u>Key domains assessed</u> | <u>Key limitations and considerations</u> |
| --- | --- | --- | --- | --- |
|  | of nutrition related risks. | concerning other nutritional issues, followed by an action plan. | nutrition-related risks | screening tools. Initial validation was limited to secure/ forensic settings. |
| **NutriMental Screener (37)** | Developed for use in adult MH settings to identify a range of nutrition related risks. | 10 items: nine nutrition-risk items and one item assessing desire for dietetic support. | BMI, weight change, appetite/eating behaviour changes, medication-related changes, disordered eating, constipation, food insecurity, nutrition-related health conditions, desire for specialist nutritional support | Developed specifically for MH services. Preliminary feasibility and validity testing was not undertaken within the UK, further validation is required and applicability to LD populations has not been established. |
| **Approaches to Schizophrenia Communication–Self-Report checklist (ASC-SR) (38)** | Identifies antipsychotic medication side effects, including those that are nutrition-related. | 17 patient-reported items, considered individually rather than combined into a total score | Antipsychotic-related adverse effects, including nutrition-relevant effects such as appetite, weight gain and gastrointestinal symptoms. | Developed as a communication checklist rather than a nutritional screening tool; initial evidence derived from a multicentre pilot evaluating clinical utility. |
| **Adapted Nutrition Risk Score (27)** | General nutrition risk screening adapted for MH populations. | 5 components adapted for use with psychiatric inpatients | BMI, unintentional weight loss, appetite, ability to eat or retain food and clinical or medical stress factors. | Primarily assesses risk of undernutrition. Limited validation of MH-specific adaptations. |

Australian research identifies unclear role responsibilities and a lack of appropriate tools as barriers to nutritional screening in MH settings (28), while UK evidence in MH/LD remains limited. This study therefore explored dietetic perspectives on nutritional screening in UK MH/LD settings to inform a British Dietetic Association (BDA) Mental Health Specialist Group (MHSG) Nutritional Screening in MH position statement.

Objectives were to:

- Identify which nutritional screening tools are used in MH/LD inpatient settings
- Explore how these tools are implemented in practice
- Examine dietitians’ views on the usefulness of these tools
- Identify perceived barriers and facilitators to effective screening
- Gather recommendations to improve screening in these settings.

## Methods

### Study design

A cross-sectional convergent mixed-methods survey collected quantitative and qualitative data concurrently and in parallel (40), with both contributing equally to study objectives. This enabled description of nutritional screening practices alongside exploration of dietitians’ experiences and perspectives, reflecting the study’s descriptive and exploratory objectives.

### Data Collection

Data was collected anonymously via Jisc Online Surveys over 63 days (April-June 2025). The novel 22-item survey, informed by the study aims and a targeted literature review, included 19 closed and three open-ended questions across five domains: demographics and professional background (3 items); screening tool use, implementation and perceived effectiveness (11 items); implementation barriers (3 items); facilitators and recommendations (2 items); and dietitians’ roles in screening (3 items) (Supplementary File 1). Pilot testing with 2 MH dietitians led to minor revisions for clarity and relevance. Ethical approval was granted by Teesside University (2025Mar26544).

### Participants and Recruitment

Convenience sampling recruited UK-registered dietitians with current or recent experience of nutritional screening in MH/LD inpatient settings, where screening procedures were considered more likely to be developed (as opposed to community services). Eating disorder services were excluded due to differences in nutritional risk identification and management.

Participants were recruited via the BDA MHSG, selected for its access to a national network of ∼1000 potentially relevant professionals. Though, as members span many specialisms, the proportion meeting eligibility criteria was unknown and a response rate could not be calculated. A single member mailer was distributed containing a participant information sheet and survey link, no reminders were sent. Participation was voluntary, with informed consent implied by proceeding to the survey questions. Responses were anonymous, with no personally identifiable information collected.

### Data Analysis

Following exportation from Jisc Online Surveys, quantitative data was analysed descriptively in Microsoft Excel using frequencies and percentages.

Qualitative data was analysed using Braun and Clarke’s reflexive thematic analysis (41) (pp. 35–36), combining deductive coding informed by study objectives and survey domains with inductive coding of concepts identified within data. Initial coding was undertaken by the student researcher and was primarily semantic. Developing codes and themes were discussed during supervision, where alternative interpretations, relationships between codes and developing theme meaning were considered, informed by supervisors greater experience of the clinical specialism. Following completion of the student project, the supervisory team further refined themes by revisiting data, considering theme coherence and distinctiveness, and developing initially descriptive themes towards more interpretive accounts. NVivo supported data organisation and theme development.

Consistent with a convergent design, quantitative and qualitative data were initially analysed separately and subsequently merged during interpretation (36). Integration involved the research team considering the quantitative results and qualitative themes together, with both informing the overall interpretation of findings.

### Researcher reflexivity

The student researcher was completing a Master’s pre-registration dietetics programme, supervised by three experienced MH dietitians spanning clinical and academic roles. Their professional backgrounds provided contextual insight but may have influenced interpretation through existing knowledge and assumptions. Reflexivity was supported through reflective discussion of developing interpretations and alternative perspectives.

This study is reported in accordance with the Good Reporting of A Mixed Methods Study (GRAMMS) criteria (42) (Supplementary File 2).

## Results

Of 48 responses, 47 were included; one was excluded due to substantial missing data. As questions were optional, response numbers varied by item.

Participant demographics are presented first, followed by integrated quantitative and qualitative findings organised thematically.

### Demographics

Almost half of participants (46%, n=21) held dietetic registration for over 10 years followed by 26% (n=12) for 7–10 years, 13% (n=6) 4–6 years, 9% (n=4) 1–3 years, and 7% (n=3) less than 1 year. Most (53%, n=25) reported ≥4 years of experience in MH/LD (Table 2).

**Table 2:** Distribution of participants by years of experience within each practice setting.

|  | Number of years within current MH/LD specialism |  |  |  |  |  |
| --- | --- | --- | --- | --- | --- | --- |
|  | <1 year | 1-3 years | 4-6 years | 7-10 years | > 10 years | Total |
| **Working-age adult inpatient mental health** | 2 | 7 | 2 | 3 | 6 | **20** |
| **Adult forensic or long-stay rehabilitation inpatient services** | 1 | 3 | 3 | 1 | 2 | **10** |
| **Older adult inpatient mental health** | 2 | 3 | 1 | 0 | 1 | **7** |
| **Children and young people's mental health inpatient services** | 0 | 2 | 1 | 1 | 0 | **4** |
| **Adult learning disabilities inpatient services** | 0 | 1 | 0 | 0 | 0 | **1** |
| **Other** | 0 | 1 | 0 | 1 | 3 | **5** |
| **Total** | **5** | **17** | **7** | **6** | **12** | **47** |

Most participants worked in working-age adult inpatient MH (43%, n=20), with representation across forensics, older adult, child and adolescent, and adult LD inpatient settings (Table 2). A further 11% (n=5) selected “Other”, describing roles spanning multiple services or including community elements.

### Theme 1 - Doing the screening, doubting the value

This theme describes a disconnect between screening implementation and its perceived impact upon patient care.

Nutritional screening practices were longstanding with 43% (n=19) introducing screening >10 years ago, 32% (n=14) within 4–10 years, and 18% (n=8) within the past 1–3 years. Seven percent (n=3) had not yet implemented screening within their service. Over half of participants (57%, n=27) reported at least 75% of patients in their setting were screened, with a further 19% (n=9) reporting screening coverage between 51–75%. Thirteen percent (n=6) believed fewer than 50% of patients were screened, including 4% (n=2) who estimated coverage below 25%. The remaining 11% (n=5) were unsure. Nutritional screening policy and auditing processes were commonplace, reported by 87% and 74% of participants respectively.

Few participants (23%, n=11) believed nutritional risk was adequately captured, with 57% (n=27) disagreeing and 19% (n=9) unsure.

**Table 3:** Quantitative results presented under theme 1, detailing screening practices.

|  |  |  |  |
| --- | --- | --- | --- |
| Nutritional screening practice implementation (n=44) | **>10 years ago** | **43%** | **n=19** |
|  | 4-10 years ago | 32% | n=14 |
|  | 1-3 years ago | 18% | n=8 |
|  | Screening not yet implemented | 7% | n=3 |
| Estimated percentage of patients screened (n=47) | **More than 75%** | **57%** | **n=27** |
|  | 51-75% | 19% | n=9 |
|  | 25-50 | 9% | n=4 |
|  | <25% | 4% | n=2 |
|  | Unsure | 11% | n=5 |
| Nutritional screening policy in place (n=46) | **Yes** | **87%** | **n=40** |
|  | No | 4% | n=2 |
|  | Unsure | 9% | n=4 |
| Auditing process in place (n=46) | **Yes** | **74%** | **n=34** |
|  | No | 13% | n=6 |
|  | Unsure | 13% | n=6 |
| Nutritional screening risk adequately captured (n=41) | Yes | 23% | n=11 |
|  | **No** | **57%** | **n=27** |
|  | Unsure | 19% | n=9 |

Free-text responses corroborated the idea of established practices, yet a failure to translate into improved care.

> “It is more of a tick box exercise. Completing the MUST is supposed to be part of regular auditing by the matrons but the actions are not monitored. So is doing a MUST really improving care. It should be about identification and action.” [P7:MUST].
>
> “Nutrition screening seems to be just another tick box exercise for ward staff.” [P17:SANSI].

Specifically related to MUST, participants suggested Step 3; ‘Acute disease effect score’, was not applicable or well understood within their setting.

> “Acute disease score does not adequately identify the poor intake that can be associated with mental health conditions” [P31:MUST].
>
> “Poor understanding of acute disease effect in mental health setting” [P23:MUST].

### Theme 2 - Beyond the tool: Structural barriers to meaningful screening

This theme reflects structural barriers that limit the impact of nutritional screening beyond the choice of tool itself (Figure 1).

**Figure 1:**
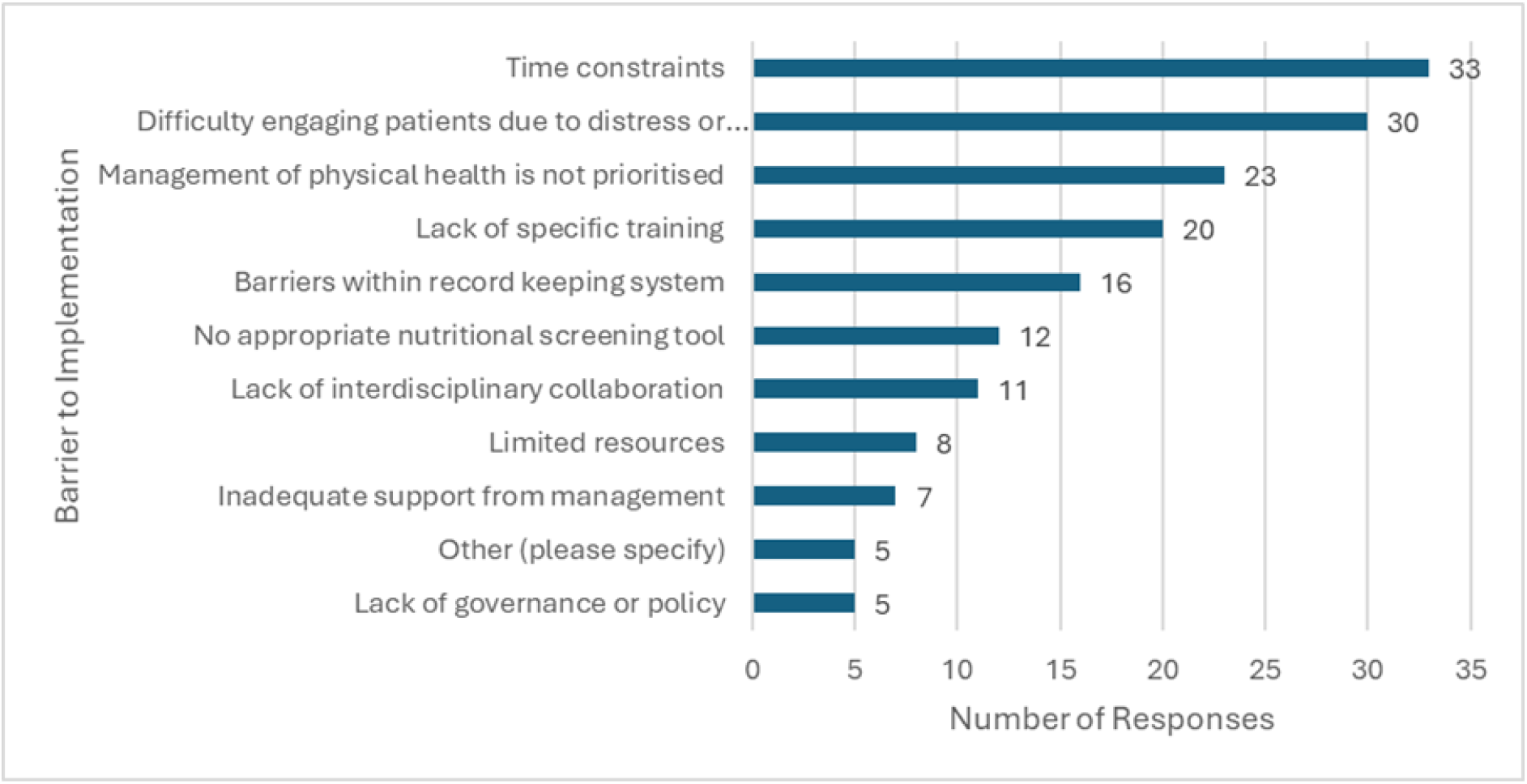
Barriers to nutritional screening implementation and effectiveness in mental health and learning disability settings.

Whilst some barriers were contextualised to MH/LD (e.g., difficulty engaging patients due to distress or mental state (64%, n=30) a lack of prioritisation of physical health (49%, n=23) and lack of appropriate screening tools (26%, n=12)), time constraint was the most frequently selected barrier; 70% (n=33). Additional barriers included a lack of training (43%, n=20), issues with record-keeping systems (34%, n=16), limited interdisciplinary collaboration (23%, n=11), and resource limitations (17%, n=8).

Almost half of respondents indicated minimal organisational support with regards to screening implementation (45%, n=21), followed by 26% reporting moderate support (n=12), 17% good support (n=8), and 9% full support (n=4). Four percent reported no organisational support for screening implementation (n=2).

Free text comments emphasised screening effectiveness relying upon wider system functionality and infrastructure.

> “The tools are only as effective as the systems in place around them… without appropriate care pathways / actions following up nutrition screening outcomes, triaging, staff buy-in & time, patient engagement with the process / questions, medical record systems (eg Rio, Paris), etc, they are less effective.” [P9:No tool].

Digital capability was often deemed essential for improving screening effectiveness.

> “Electronic tool which does maths for them!” [P32:MUST].
>
> “We have a new EPR and the tool we are using makes the subjective scoring compulsory which [h]as increased accuracy” [P43:Bespoke tool].

Digital integration also dictated which tool was used.

> “The MUST is attached to our RIO electronic notes which is why we use it” [P7:MUST].

### Sub theme: The role of others

Most participants (49%, n=23) reported a secondary role in nutritional screening, with only 15% (n=7) actively screening. Some had limited (2%, n=1) or no (2%, n=1) involvement. Many reported leadership responsibilities for screening, either as a primary lead (34%, n=16) or as a shared responsibility (40%, n=19).

Most participants identified registered nurses as completing nutritional screening (78%, n=36), followed by nursing assistants (57%, n=26) and dietitians (24%, n=11). Other staff groups included healthcare assistants, dietetic support workers, support workers, exercise therapists, doctors, and ward managers.

Perceived importance of nutritional screening among non-dietetic colleagues was modest: 47% (n=22) rated it moderate, 38% (n=18) low and 11% (n=5) very low. Only one respondent (2%) rated it high, with none rating it very high.

**Table 4:**
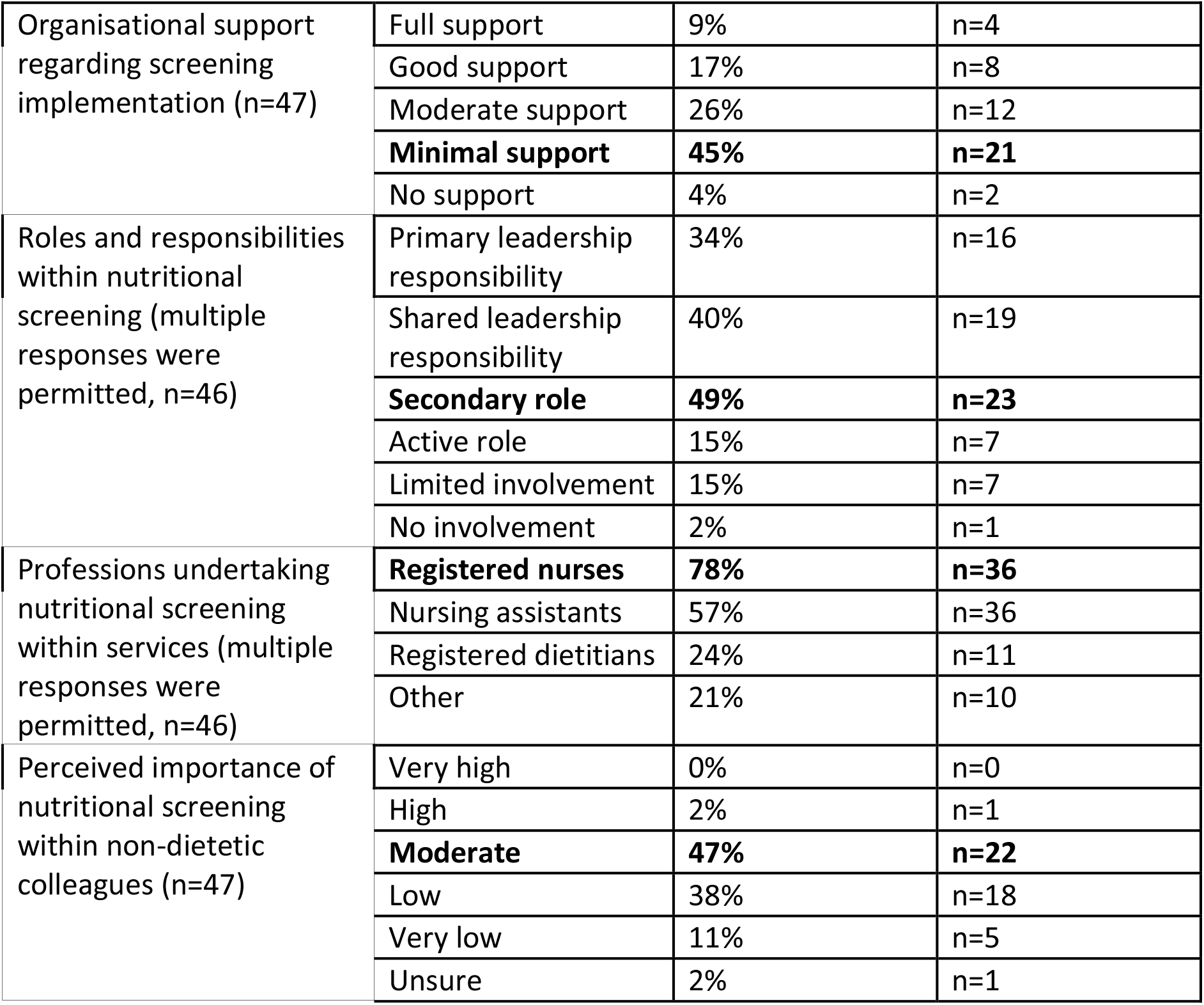
Quantitative results presented under theme 2, detailing roles and responsibilities within screening.

Free-text comments highlighted that human error can impact screening accuracy.

> “We get errors through miscalculation of BMI… very rarely get % weight loss.” [P43:Bespoke].
>
> “Scoring is not correct and staff find % weight loss hard” [P41:MUST].

### Theme 3 - Redefining screening scope

This theme highlights participants’ expectation that nutritional screening tools should capture broader nutritional risks experienced in MH/LD.

MUST was used by most respondents (66% (n=31)) followed by bespoke tools (15% (n=7)) and SANSI (9%, n=4)). Other responses included the Mini Nutritional Assessment (2%, n=1) and Nutritional-Risk Screening 2002 (NRS-2002) (2%, n=1). A further 4% (n=2) selected “Other”, specifying WAASP and PYMS (Figure 3) and 4% (n=2) did not use a screening tool.

**Figure 2:**
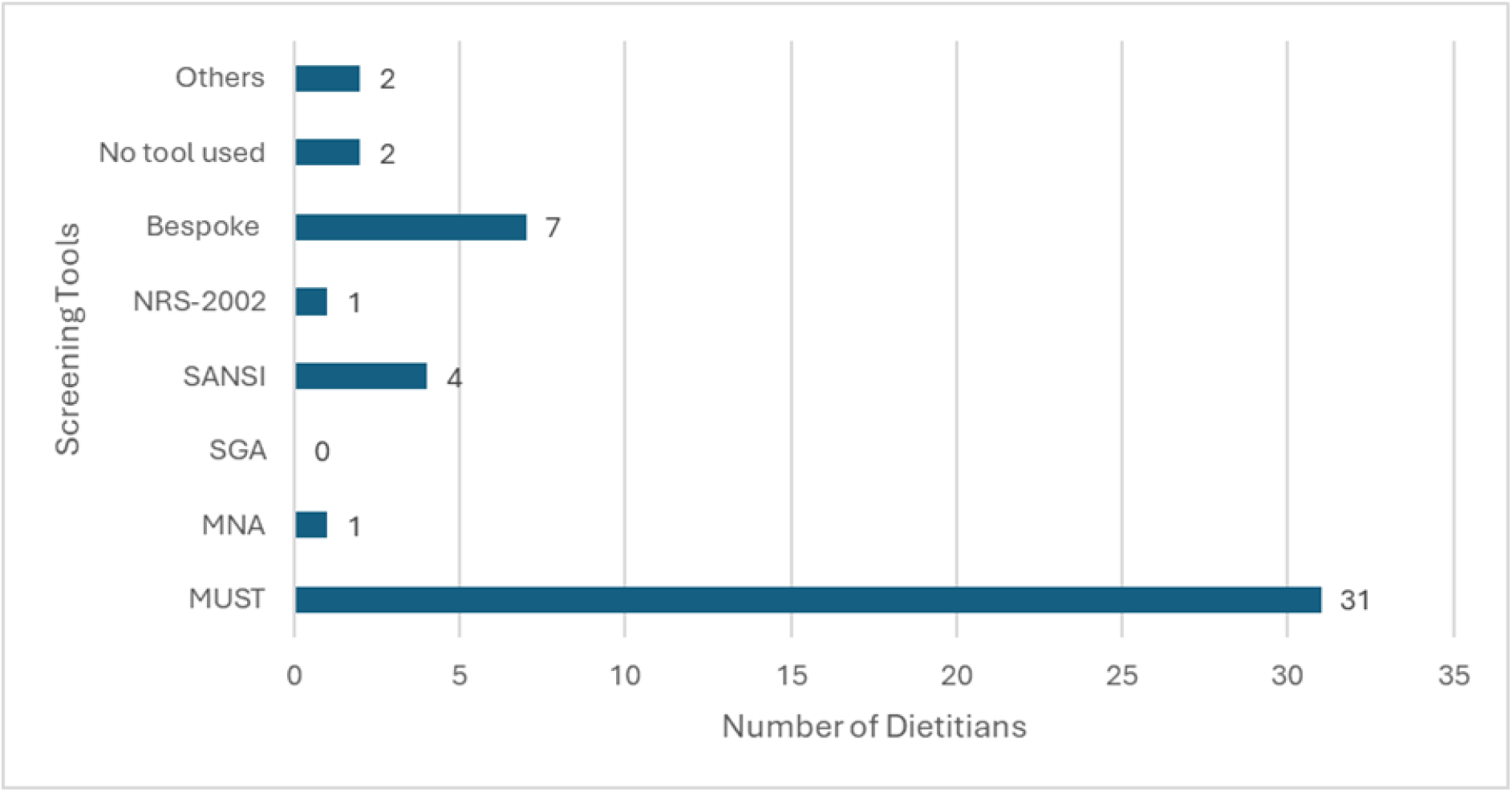
Frequency of Nutritional Screening Tool Use (n = 47)

**Figure 3:**
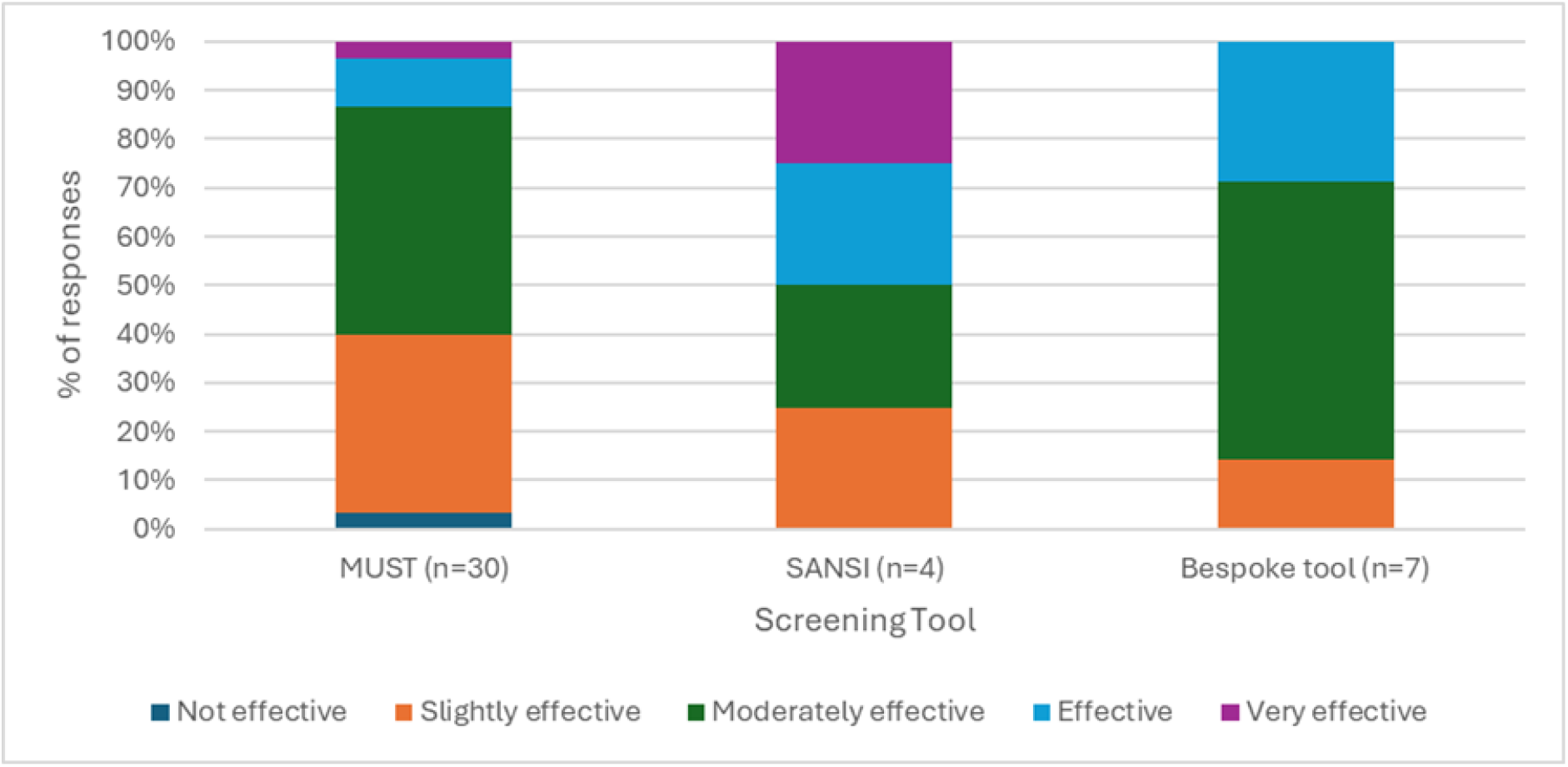
Perceived effectiveness of screening tools (n = 41)

Bespoke tools were rated most positively while MUST was generally seen as moderately (45%, n=14) or slightly effective (36%, n=11). Views on SANSI varied widely (Figure 4).

Free text comments highlighted how tools were likely to identify undernutrition but neglect other commonly observed nutritional risks.

> “Good at identifying malnutrition but not for other nutritional problems this patient group may experience, e.g. disordered eating, weight gain associated with anti-psychotics, cardiovascular disease.” [P42:MUST].
>
> “Not specific to identify cardiometabolic risk factors… not specific to explore food insecurity issues.” [P45:MUST].
>
> “Current tools used only take into account physical reasons… not mental/psychiatric reasons.” [P13: MUST:WASP].

Where SANSI was used, it was perceived as overly sensitive, resulting in widespread ‘high-risk’ categorisation.

> “SANSI effectively identifies nutrition risks; however, its lack of specificity is challenging. Since 80% of Forensic Mental Health Inpatient service users live with obesity, SANSI classifies almost all of them as high nutrition risk.” [P16:SANSI].
>
> “In a service where overweight and obesity are highly prevalent and diabetes is common, most service users score ‘high’. It’s correct as they’re at risk, but it would be helpful to differentiate further e.g. high & very high.” [P17:SANSI].

Another challenge related to screening scope was the need for a tool flexible enough to accommodate different patient groups with distinct nutritional needs.

> “Challenging for Trusts to agree on a tool to be used across all settings e.g. want to keep MUST in older peoples services but SANSI better suited for working age adults.” [P43:MUST].

## Discussion

Vast health inequalities exist within MH and LD populations with preventable diet-related conditions contributing substantially to excess mortality (1–9). Despite the disproportionate presence of undernutrition, obesity and several nutrition-related or nutrition-impacting conditions, nutritional screening methods and their validation in these settings is under-researched (29). To our knowledge, this is the first study to explore dietitians’ perspectives on nutritional screening practices within UK MH/LD inpatient settings. Findings indicated established screening practices that are audited and policy-driven, however, participants expressed dissatisfaction with existing tools, citing an inability to capture the multifactorial nature of nutritional risk, alongside broader systemic and procedural barriers.

Longstanding concerns about the limited scope of nutritional screening beyond undernutrition have driven the development of MH specific tools (35,37). Concerns are warranted given the inherently multifactorial nature of nutritional risk in MH/LD populations, where metabolic, behavioural, pharmacological and environmental factors often intersect with risk of over, under or imbalanced nutrition. However, key principles of effective nutritional screening including speed, ease of use and suitability for non-expert users (43), could be difficult to uphold if expanding screening to a wide range of nutritional risks. That said, the 10-item NutriMental screener (37), addresses the multifactorial nature of nutritional risk within MH with positive preliminary feasibility testing (39). Similarly, SANSI captures a range of nutritional risks and training is reportedly achievable in under 10 minutes (35). While further testing of the NutriMental screener is planned (39), evidence for SANSI is to our knowledge, limited to its development study in secure inpatient services and one subsequent single site study reporting positive acceptability across MH/LD wards (36). Further research into its validity, feasibility and clinical utility would support more confident endorsement of wider implementation.

Most study participants utilised MUST, reflecting its widespread clinical use and inclusion in national guidelines (31–33). However, it was most commonly rated as only slightly or moderately effective, with qualitative findings highlighting concerns about its ability to capture the breadth of nutritional risk in MH/LD. This may suggest that use reflects organisational drivers and limited alternatives rather than clinical confidence. Alongside concerns that MUST does not capture nutritional risks beyond undernutrition, Step 3 (‘acute disease effect’) was often considered irrelevant or poorly understood. Early MUST guidance (30) (p.54) defines Step 3 as identifying individuals with, or likely to have, little or no nutritional intake for ≥5 days due to acute illness using exclusively physical health examples (e.g., major injury, burns, stroke). However, later guidance (44) (p.7) extends its application to acute psychological conditions resulting in minimal nutritional intake. Perceived limitations may therefore partly reflect interpretation of predominantly physical health focused guidance rather than the tool itself. Greater inclusion of MH/LD examples, such as food refusal during acute mental illness, may aid application.

Some participants used bespoke, locally developed screening tools, which were generally rated more positively than validated options, potentially reflecting familiarity or involvement in their development. While these may address perceived limitations of existing tools, unpublished local tools lack robust validation and limit comparability across settings, hindering development of a coherent evidence base (45). Efforts may be better directed towards strengthening confidence in existing validated MH-specific tools (e.g., SANSI) and/or advocating for better inclusion of MH considerations within MUST. This would support comparability, reduce duplication, and enable teams to draw on guidance, training and implementation support afforded by more established tools; areas deemed important within this study.

Beyond concerns about the tools themselves, participants identified systemic barriers to effective screening. Despite generally well-established practices, screening was often described as procedural, with limited confidence that it meaningfully influenced care, highlighting the need for stronger integration within care pathways. This finding reflects early literature describing malnutrition screening in MH as compliance-driven rather than action-oriented (27), and national guidance on improving physical health in SMI which emphasises the principle of “don’t just screen, intervene” (46). Improved integration of screening within care pathways may also help address concerns regarding SANSI’s over-sensitivity within this study. In settings where nutritional risk is commonplace, widespread identification of nutritional risk may be accurate, but presents challenges for resource-limited services and may dilute perceived importance of risk. A stratified risk output linked to proportionate, clearly defined care pathways is needed. Existing screening-led pathways show promise (36) but largely focus on weight management, with limited evidence of effective implementation across broader nutritional risks in MH/LD settings.

This study identified limited organisational support for screening, including poor prioritisation of physical health and few strategies to address challenges engaging patients in screening, reflecting barriers reported previously (47). This emphasises the need for screening tools that can be implemented pragmatically in MH/LD settings if comprehensive patient history or anthropometric measurements are not feasible. National guidance (48) recommends reasonable adjustments to physical health checks, including accessible communication, additional time, carer involvement, and trauma-informed, sensory-aware care, which may help address engagement barriers.

Aligned with wider evidence (33), time was the most frequently reported barrier to effective screening, while qualitative findings highlighted the influence of digital integration for effective screening. This emphasises the importance of organisational support, as screening tool selection, time allocation and electronic record infrastructure are likely to be determined at service or regional level and sit largely beyond individual clinicians’ control.

Dietitians were also unlikely to directly undertake nutritional screening, instead holding a predominantly secondary role. Improvements are therefore unlikely to be achieved by dietitians or organisational leadership alone. Engagement of those who routinely undertake screening is also needed to promote multiprofessional ownership and avoid a purely top-down approach (49).

## Recommendations

Improving the identification and management of nutritional risk in MH/LD settings is essential to reducing health inequalities in these populations. The BDA MHSG is well-placed to consider the following recommendations when developing a position statement to support its members in achieving this aim.

1. Nutritional screening using an appropriate validated tool is strongly recommended across all MH/LD inpatient settings. Screening processes should include clear actions proportionate to the level and type of nutritional risk identified.
2. The MHSG should collaborate with MUST developers to strengthen guidance for MH/LD settings. Given MUST’s widespread use, clearer guidance and MH/LD-specific examples may improve its applicability and perceived effectiveness.
3. Where limitations with MUST are identified, MH-specific tools such as SANSI and NutriMental may offer alternatives. However, adoption should consider digital compatibility and the more established training and implementation infrastructure supporting MUST.
4. Local bespoke tools should be used with caution due to development and validation demands and reduced comparability across settings. Where existing tools do not meet population-specific needs, regional or national collaboration is encouraged to minimise duplication.
5. Frontline staff should be supported to implement reasonable adjustments and pragmatic approaches to risk identification when MH/LD contextualities may impact screening e.g., fluctuating mental state or communication differences.
6. Organisational leadership and multiprofessional engagement should be prioritised to promote successful improvements to screening practices.
7. Maximising digital integration and functionality should be prioritised within any nutritional screening improvement initiative.
8. Dietitians should maintain up-to-date knowledge and support further research into the feasibility, validity and implementation of MH-specific nutritional screening tools, including SANSI and the NutriMental Screener.
9. Dietitians should support research into the wider organisational and implementation factors influencing effective nutritional screening and subsequent care in MH/LD settings.
10. A national audit of screening implementation may be a reliable and feasible approach to determine success of screening implementation within MH/LD settings.

## Limitations

This study did not include community MH/LD settings, and LD and child and adolescent MH inpatient services were underrepresented, limiting transferability beyond adult MH inpatient settings. As some challenges identified in the present study may reflect experiences in other settings, further research should explore screening practices and needs across community settings and those underrepresented in the present study. Additionally, screening practices were explored solely from the perspective of dietitians and relied on subjective assessments of effectiveness.

The absence of comprehensive workforce data meant the proportion of UK MH/LD inpatient dietitians represented was unknown, and practices in many services likely remain undocumented. Recruitment through a dietetic specialist group also favoured settings with dietetic provision. Many MH/LD inpatient settings do not have dedicated dietetic provision and screening practices are likely less developed than those represented in this study.

## Conclusion

MH/LD populations experience substantial nutrition-related health inequalities, highlighting the need to strengthen nutritional screening. This study identified no clear preferred tool for MH/LD inpatient settings, with limitations reported across current approaches. Effective screening requires more than selecting a valid and feasible tool. It must be supported by organisational leadership with systems integration, multiprofessional ownership and clear pathways that translate identified nutritional risk into meaningful care.

## Data Availability

All data produced in the present study are available upon reasonable request to the authors

## Acknowledgments

The authors would like to thank the dietitians working in mental health and learning disability inpatient services who participated in this study.

## Conflicts of interest

There are no conflicts of interest to declare.

Ethical approval was granted by Teesside University (2025Mar26544).

## Notes

### Competing Interest Statement

The authors have declared no competing interest.

### Author Declarations

Ethical approval was granted by Teesside University, School of Health and Life Sciences Ethics Committee (2025Mar26544).

