## Supplementary file 1 for "Nutritional screening in mental health and learning disability inpatient services: Dietitians’ perspectives on practices, barriers and tool suitability"

### Supplementary File 1. Survey Questions

#### Section 1: Demographics and Professional Background

Please answer the following questions about your current role and experience working in inpatient mental health and/or learning disability settings. Your answers will help us understand the context of your screening practices.

1. How many years have you worked as a registered dietitian?

- <1 year
- 1–3 years
- 4–6 years
- 7–10 years
- > 10 years

2. How many years have you worked in a mental health or learning disability setting?

- <1 year
- 1–3 years
- 4–6 years
- 7–10 years
- > 10 years

3. **Please select the mental health or learning disability setting in which you are predominantly based:**
*If your role covers more than one specialism, please select the one you are predominantly based within. If your specialism is not listed but meets the inclusion criteria, please choose the option that best fits your role or use the 'other' selection.*

- Working age adult inpatient mental health
- Older adult inpatient mental health
- Adult learning disabilities inpatient services
- Adult forensic or long-stay rehabilitation inpatient services
- Children and young people's mental health inpatient services
- Children and young people's learning disability inpatient services
- Children and young people's forensic or long-stay rehabilitation inpatient services
- Other (please specify)

#### Section 2: Use and Perceived Effectiveness of Nutritional Screening Tools

The following questions explore your use of nutritional screening tools within inpatient mental health and/or learning disability settings and your views on their suitability and effectiveness.

4. What nutritional screening tools do you commonly use in your practice with mental health patients? (Select all that apply)

- ☐ Malnutrition Universal Screening Tool (MUST)
- ☐ Mini Nutritional Assessment (MNA)
- ☐ Subjective Global Assessment (SGA)
- ☐ St. Andrew's Nutrition Screening Instrument (SANSI)
- ☐ Nutritional Risk Screening 2002 (NRS-2002)
- ☐ Bespoke trust-wide tool
- ☐ No tool used
- ☐ Other (please specify)

5. When did your service first implement nutritional screening for mental health patients?

- ○ Less than 1 year ago
- ○ 1–3 years ago
- ○ 4–10 years ago
- ○ More than 10 years ago
- ○ Not Implemented

6. How effective do you find these tools in assessing the nutritional risk of mental health patients?

- ○ Not effective
- ○ Slightly effective
- ○ Moderately effective
- ○ Effective
- ○ Very effective

7. Please explain why you rated the effectiveness of these tools as you did.

8. In your opinion, do current nutritional screening tools adequately capture the unique nutritional risks associated with your patient group?

- ○ Yes
- ○ No
- ○ Not sure

9. In your opinion, how frequently do nutritional screening tools fail to identify nutritional risks specific to mental health patients?

- ◯ Never
- ◯ Rarely
- ◯ Sometimes
- ◯ Often
- ◯ Always
- ◯ N/A - not using

10. Please (if applicable), explain why you think current nutritional screening tools do not adequately capture the risks.

11. Does your organisation have policies related to nutritional screening (e.g., screening within a specific timeframe after admission)?

- ◯ Yes
- ◯ No
- ◯ Not sure

12. Is nutritional screening audited in your organisation?

- ◯ Yes
- ◯ No
- ◯ Not sure

13. Approximately, what proportion of patients in your care are screened for nutritional risk?

- ◯ Less than 25%
- ◯ 25–50%
- ◯ 51–75%
- ◯ More than 75%
- ◯ Unsure

14. Who typically undertakes nutritional screening in your setting? (Select all that apply)

- ☐ Registered Dietitian
- ☐ Registered Nurse
- ☐ Nursing Assistant
- ☐ Other (please specify)

#### Section 3: Barriers to Implementation

The following questions aim to explore the key challenges you experience when using nutritional screening tools within inpatient mental health and/or learning disability settings.

15. How often do day-to-day logistical challenges (e.g., staff shortages, lack of training) prevent timely and effective nutritional screening?

- ◯ Never
- ◯ Rarely
- ◯ Sometimes
- ◯ Often
- ◯ Always

16. To what extent do you feel that organisational support is available to implement nutritional screening effectively?

- ◯ No support
- ◯ Minimal support
- ◯ Moderate support
- ◯ Good support
- ◯ Full support

17. What are the most significant barriers to the effective implementation of nutritional screening in your practice setting? (Select all that apply)

- ☐ Time constraints
- ☐ Lack of specific training
- ☐ Limited resources
- ☐ Inadequate support from management
- ☐ Lack of interdisciplinary collaboration
- ☐ Difficulty engaging patients due to distress or acute mental health symptoms
- ☐ No appropriate nutritional screening tool
- ☐ Barriers within record keeping system
- ☐ Lack of governance or policy
- ☐ Management of physical health is not prioritised
- ☐ Other (please specify)

#### Section 4: Facilitators and Recommendations

The following questions aim to explore what would help improve the use and effectiveness of nutritional screening tools in inpatient mental health and/or learning disability settings. Your insights will help identify opportunities for better practice and support.

18. Which factors or resources would most improve the effectiveness of nutritional screening in your setting? (Select all that apply)

- [ ] Staff training on nutritional needs within your patient group
- [ ] Staff training on nutritional screening tools
- [ ] Access to a suitable or mental health-specific nutritional screening tool
- [ ] Interdisciplinary collaboration
- [ ] Time allocated specifically for screening
- [ ] Improved functionality within record keeping system
- [ ] Improved governance e.g. nutritional screening policy and audit
- [ ] Other (please specify)

19. What (if any) additional features or adaptations would make existing nutritional screening tools more suitable for your patient group?

#### Section 5: Role of Dietitians in Screening Practices

This section focuses on your involvement and views regarding the role of dietitians in nutritional screening within inpatient mental health and/or learning disability settings.

20. How would you best describe your role in supporting nutritional screening practices within your practice setting? (select all that apply)

- [ ] Primary leadership responsibility e.g. implementing and embedding process
- [ ] Shared leadership responsibility with other healthcare professionals
- [ ] Secondary role e.g. promoting implementation
- [ ] Active role e.g. physically undertaking nutritional screening
- [ ] Limited involvement
- [ ] Not involved
- [ ] Unsure

21. How frequently do you collaborate with other healthcare professionals (e.g., nurses and physicians) in conducting nutritional screenings for mental health patients?

- ○ Never
- ○ Rarely
- ○ Sometimes
- ○ Often
- ○ Always

22. In your experience, what is the level of understanding among non-dietitian healthcare professionals regarding the importance of nutritional screening in mental health?

- Very low
- Low
- Moderate
- High
- Very high
- Unsure
