## Supplementary file 2 for "Nutritional screening in mental health and learning disability inpatient services: Dietitians’ perspectives on practices, barriers and tool suitability"

**Supplementary File 2. Good Reporting of A Mixed Methods Study (GRAMMS) checklist**

O'Cathain A, Murphy E, Nicholl J. The quality of mixed methods studies in health services research. J Health Serv Res Policy. 2008 Apr;13(2):92-8. doi: 10.1258/jhsrp.2007.007074. PMID: 18416914.

| GRAMMS criteria | Application | Manuscript section |
|---|---|---|
| Describe the justification for using a mixed methods approach to the research question | A convergent mixed-methods design enabled description of nutritional screening practices alongside exploration of dietitians experiences and perspectives, reflecting the descriptive and exploratory study objectives. | Methods – study design |
| Describe the design in terms of the purpose, priority and sequence of methods | Quantitative and qualitative data were collected concurrently and in parallel, with both methods contributing equally to addressing the study objectives. | Methods – study design |
| Describe each method in terms of sampling, data collection and analysis | The same sampling approach was used in both components. Quantitative and qualitative data were collected concurrently through closed- and open-ended survey questions and analysed separately using descriptive statistics and reflexive thematic analysis, respectively. | Methods – study design, data collection, participants and recruitment, analysis |
| Describe where integration has occurred, how it has occurred and who has participated in it | Integration occurred during interpretation, with the research team considering quantitative results and qualitative themes together to inform the overall interpretation. | Methods – data analysis |

| | | |
|---|---|---|
| Describe any limitation of one method associated with the presence of the other method | No specific limitations arising from the presence of one method alongside the other were identified. Other limitations are reported. | Limitations |
| Describe any insights gained from mixing or integrating methods | Integrated interpretation identified several complementary insights, including differences between established screening practices and their perceived effectiveness and clinical meaningfulness. | Discussion |
